# Modeling the Covid-19 Pandemic Response of the US States

**DOI:** 10.1101/2020.06.24.20138982

**Authors:** Georgios Neofotistos, Efthimios Kaxiras

## Abstract

**Background:** The United States of America (USA) has been the country worst affected, in absolute terms, by the Covid-19 pandemic. The country comprises 50 states under a federal system. The impact of the pandemic has resulted in different responses at the state level, which are driven by differing intervention policies, demographics, connectedness and other factors. Understanding the dynamics of the Covid-19 pandemic at the state level is essential in predicting its future evolution.

**Objective:** Our objective is to identify and characterize multiple waves of the pandemic by analyzing the reported infected population curve in each of the 50 US states. Based on the intensity of the waves, characterized by declining, stationary, or increasing strengths, each state’s response can be inferred and quantified.

**Methods:** We apply a recently proposed multiple-wave model to fit the infected population data for each state in USA, and use the proposed Pandemic Response Index to quantify their response to the Covid-19 pandemic.

**Results:** We have analyzed reported infected cases from each one of the 50 USA states and the District of Columbia, based on the multiple-wave model, and present the relevant parameters. Multiple waves have been identified and this model is found to describe the data better. Each of the states can be classified into one of three distinct classes characterized by declining, increasing, or stationary strength of the waves following the initial one. The effectiveness of intervention measures can be inferred by the peak intensities of the waves, and states with similar population characteristics can be directly compared. We estimate how much lower the number of infections might have been, if early and strict intervention measures had been imposed to stop the disease spread at the first wave, as was the case for certain states. Based on our model’s results, we compute the value of the Pandemic Response Index, a recently introduced metric for quantifying in an objective manner the response to the pandemic.

**Conclusions:** Our results reveal a series of epidemic waves, characterizing USA’s pandemic response at the state level, and also infer to what extent the imposition of early intervention measures could have had on the spread and impact of the disease. As of June 11, 2020, only 19 states and the District of Columbia (40% of the total) clearly exhibit declining trends in the numbers of reported infected cases, while 13 states exhibit stationary and 18 states increasing trends in the numbers of reported cases.

## Introduction

The Covid-19 pandemic has generated a plethora of studies aspiring to understand the dynamics of the disease spread and predict its future evolution (see, for example, [1-9]). Different types of models can be assumed to describe this dynamic evolution. In studying past epidemics, scientists have systematically applied “random mixing” models which assume that an infectious individual may spread the disease to any susceptible member of the population, as originally proposed by Kermack and McKendrick [10]. Recent approaches consider the effects of mobility and contacts in networks [11], epidemic waves attributable to community networks [12], sub-epidemic modeling [13], Bayesian modeling and inference [14], agent-based simulations of social distancing measures [15], and power-law models of infectious disease spread [16], to name but a few representative examples.

Modeling the effect of the imposition of social distancing measures has been a very active area of study, as the imposition of these measures is considered to be the most effective policy for mitigating the disease [17, 18]. In the reported infections in many countries and states, there appear to exist regular features consisting of persistent, non-random, wavy behavior in the epidemic trajectory; this behavior can be linked to the effect of intervention measures, which reveals useful information for predicting the temporal evolution of the pandemic.

Agent-based simulations encompassing strong social-distancing measures show the emergence of epidemic trajectories with multiple wave structures [15], and recent research has focused on interpreting the wave features appearing on the reported incidence curves by “change points” resulting from change in the epidemiological parameters after the imposition of interventions [14] or by decomposing the epidemic trajectory in multiple waves [15].

Recent results on the introduction and spread of the virus in Arizona [19], based on sequencing viral genomes from clinical samples, collected as part of community surveillance projects and particular phylogenetic analysis of genomes, reveal a minimum of 9 distinct introductions throughout February and March 2020. Furthermore, it was shown that the first reported case of community transmission in Arizona descended from the Washington state outbreak discovered in late February 2020, but none of the observed transmission clusters were found to be epidemiologically linked to the original travel-related cases in the state, suggesting early isolation and quarantine effects. These findings corroborate the epidemic evolution of Covid-19 in multiple waves (clusters, outbreaks, sub-epidemics) as presented in [15], with several clusters of infections appearing in waves due to different spatial introductions of the disease, starting subsequent local outbreaks of infections.

As the Covid-19 pandemic appears to have been contained in Europe and in Asia, there is a serious concern about the possibility of the emergence of a “second wave” (or “surge”) of the Covid-19 pandemic, and what its impact will be (see, for example, [20, 21]). At present, there is a trove of data from different countries, states, counties, and cities that can serve to validate and put strict limits on plausible models. In this paper, based on our previous work on simulations from agent-based models and a multiple wave model, we fit reported cases data from each of the 50 USA states and the District of Columbia. The results provide a comprehensive picture of likely scenarios of how the disease evolved in the country up to June 11, 2020. These scenarios can be useful in predicting the future spread of the disease at the state level and provide insight on how the imposition of social-distancing measures can be effective in containing or slowing its spread.

Our work is based on two premises: First, that the apparent regular (wavy) features in the reported infections in many states are not random, but rather contain useful information, as their persistence and regularity across many regions and countries suggest. Second, that there is a general underlying dynamics of the spread of the disease, *in spirit* similar to the original Kermack-McKendrick model of three populations [10], the “susceptible population” *S*(*t*), the “infected/infectious population” *I*(*t*) and the “removed/recovered population” *R*(*t*), which are related by *S*(*t*) + *I*(*t*) + *R*(*t*) = *N*, where *N* is the total population. The time evolution of the *S* − *I* − *R* (SIR) populations is described by the equations:

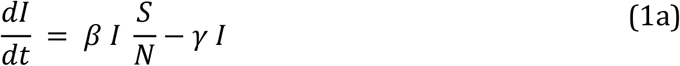

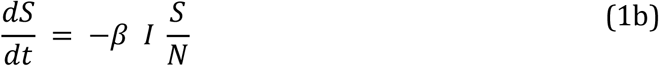

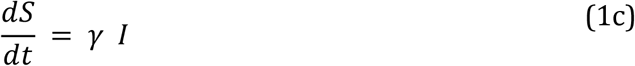

The SIR model involves two positive parameters, *β* and *γ*, with the following meaning:

- *β* describes the effective contact rate of the disease: an infected individual comes into contact with *β* other individuals per unit time (the fraction that is susceptible to contracting the disease is *S/N*);
- *γ* is the mean removal (recovery) rate, that is, 1*/γ* is the mean time during which an infected individual can pass it on before being removed from the group of the infected individuals.

However, the time evolution of the SIR populations as captured by the linear first-order differential equations of the Kermack-McKendrick model, which implicitly assumes a single peak epidemic wave, produce behavior that is much simpler than actual reported data of infections. In this paper, we fit the data with a multiple wave FSIR model with few adjustable parameters, as explained below, and use the results of the fitting to draw insights on the actual evolution of the disease in the US states.

## Methods

In order to understand the dynamics of the Covid-19 epidemic, the forced-SIR (FSIR) model was recently proposed by the authors [22] and was used to describe the evolution of Covid-19 pandemic in a representative set of countries [15]. The original FSIR model treated the evolution of the infected population as a single-peak wave, the simplest possible model, and relied on three adjustable parameters that were estimated for each country by fitting actual data. However, the single-wave assumption cannot explain the *entire* incidence curve (infected population curve). Wavy patterns with multiple local peaks are evident in the actual data, which cannot be attributed to simply random fluctuations or lower reported case numbers on weekends, due to their regularity and similarity among several countries or states in which, at first glance, were affected by the disease at different levels of severity.

In its original version FSIR applies to a single epidemic wave, in which the infected population is given by the expression

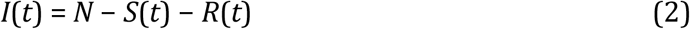

with the approximate solution given by:

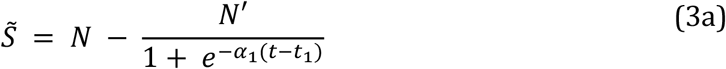

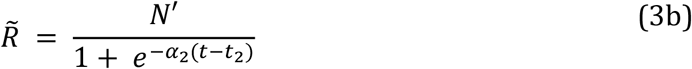

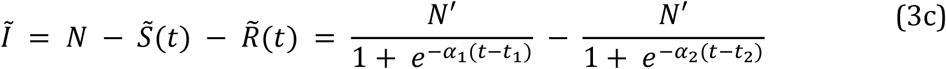

where *N*^’^, *α*_1_, *α*_2_, *t*_1_, *t*_2_ are treated as adjustable parameters, with *t*_1_ and *t*_2_ representing the times at which the 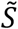 and 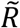 populations reach their sigmoid midpoint values, respectively. Here, we extend this model to allow for multiple waves that capture the sub-epidemics in the infected population of a country or state. For each wave *i*, the respective adjustable parameters are *N*^’*(i)*^, *α*_1_^*(i)*^, *α*_2_^*(i)*^, *t*_1_^*(i)*^, *t*_2_^*(i)*^, calculated by applying, appropriately, Eq.s (3). We apply this extended model to fit the multiple wave behavior of infected populations in all 50 USA states and the District of Columbia, as obtained from the New York Times Covid-19 data source [23], for a period ending on June 11, 2020, which corresponds to almost 150 (21.5 weeks) days from the onset of the exponential growth of reported cases in China. Population statistics for each state were obtained from the census.org data source [24]. To obtain a meaningful fit we had to consider data that show a monotonic increase at the beginning. This means that a few data points in each case were excluded, as they corresponded to sporadic reports of very few isolated cases, typically 1 to 10 in a given day, interspersed by several days of zero cases. In practice this means that the fitting begins at a certain cutoff day denoted as *t*_0_.

As in the case of the original FSIR model, in order to make the fit more robust and simpler we chose the *α*_1_ and *α*_2_ parameters to be: *α*_1_ = 0.25 (see relevant discussion in [15]) and *α*_2_ = 0.66 *α*_1_; this last relationship was established by obtaining the best fit between the approximate solution represented by Eq. (3) and the actual numerical solution of the SIR equations for parameter values relevant to the pandemic, namely *β*=0.25, *γ*=1/14. These values were not chosen merely on theoretical grounds but were estimated by fitting actual reported data. The State of Hawaii offers an ideal example because it imposed early travel restrictions and measures (individuals, both residents and visitors, arriving from out-of-state to Hawaii were subject to enhanced passenger verification process and a mandatory 14-day self-quarantine, which was also imposed to interisland travel). This imposition of early “distancing” measures resulted in a clear single-wave epidemic, which we fitted obtaining parameter values of *α*_1_ = 0.23 and *α*_2_ = 0.15: the first of these value is very close to the *α*_1_ = 0.25 value obtained in [22], whereas the second one satisfies the relation *α*_2_ = 0.652×*α*_1_, in very good agreement with the result mentioned above.

Finally, instead of using *t*_1_ and *t*_2_ for each wave as independent parameters, we elected to use as independent parameters *t*_1_ and Δ*t* = *t*_2_ − *t*_1_. To provide a numerical estimate of the value of Δ*t* we fitted actual data reported by the single-wave State of Hawaii, obtaining Δ*t* = 14 days; the same value is obtained by fitting reported data for Vermont and Montana, which also showed single-wave behavior. In order to make the multiple wave fits more robust, simpler, and systematic, we chose Δ*t* = 14 days for all four-wave fits and for states exhibiting single-wave behavior. This choice is further supported by the common mean time of 14 days before the infected individual is removed, imposed as a quarantine measure for the majority of countries around the world imposing restrictive (non-pharmaceutical) measures, and is consistent with a reported estimated median time of approximately 2 weeks from onset to clinical recovery for mild cases [25]. With *α*_1_ = 0.25, the choice Δ*t* = 14 days corresponds to the value of basic reproduction number for the Covid-19 disease R_0_=3.5 (see detailed discussion in [22]), which is very close to the average, R_0_=3.28, of values obtained by different methods [26]. For states exhibiting sub-epidemics of large intensities (peaks), we used the value Δ*t* = 21 days because these large peaks are the result of superposition of several smaller outbreaks, an apparent characteristic in states with large populations and urban centers; this larger value effectively averages the smaller waves into fewer features. By fixing the value of the parameter Δ*t*, based on plausible epidemiological characteristics, we are left with two adjustable parameters *per sub-epidemic* that can be varied to obtain the best fit to the data: the onset time *t*_1_ which corresponds to the midpoint of the sigmoid representing the decline of the susceptible population, and *N*^’^ which is a parameter representative of the number of daily cases near the peak of the infected population curve, in the given sub-epidemic. *N*_*T*_, the total number of infected. and the *N*_*T*_^*(i)*^, the total number of infected per wave *i*, can readily be obtained. The best fit here is defined in the Root-Mean-Square (RMS) sense. The model parameters have been determined by employing the Levenberg-Marquardt algorithm.

The various states responded to the pandemic in different ways. It is an interesting question to quantify their varied response and make comparisons, which may be useful for contributing to the evaluation of the policies followed and the factors underlying the pandemic dynamics. Based on the results of our model it is possible to construct an index [15], the Pandemic Response Index (PRI), and assign a value to each state depending on its response to the pandemic. To do this in an objective manner, we take three factors into consideration:

a. The total number of infections as given by the quantity *N*_*T*_ of Table I, divided by the population of the state *N*_*P*_. The range of this quantity when multiplied by a factor of 25 is between 0 and 0.5. This is a measure of the overall impact of the pandemic on the population of the state.

**TABLE I:** The values of the various parameters that enter in the multi-wave FSIR model of Eq. (3), for the 50 USA states and for the District of Columbia. The last column includes the values for the *expected* total number of cases, *N*_*T*,_ when the number of infections has dropped to near zero, and is an *extrapolated* value. *N*_*T*_^*(1)*^ is the total number of infected people in the first wave. The superscripts of the *t*_1_ and *N’* parameters represent the sub-epidemic waves. The US states are ordered according to their population size.
b. Δ*N*_*T*_, the number of cases that correspond to all the waves except for the first major one (Δ*N*_*T*_ *= N*_*T*_ *–* Δ*N*_*T*_^*(1)*^), which in some cases includes the earliest small wave (see Table II). Arguably, this number of cases could have been avoided, had the states imposed early and strict measures after the first wave of the epidemic was plainly evident, as was the case for the single-wave states. The larger this number is, the worse the performance. This number, divided by 2*N*_*T*_, lies in the range 0 to 0.5.

| USA States | $t_1^{(1)}$ | $N'^{(1)}$ | $t_1^{(2)}$ | $N'^{(2)}$ | $t_1^{(3)}$ | $N'^{(3)}$ | $t_1^{(4)}$ | $N'^{(4)}$ | $N_T^{(1)}$ | $N_T$ |
| --- | --- | --- | --- | --- | --- | --- | --- | --- | --- | --- |
| California | 29.0 | 1,999 | 50.8 | 2,385 | 69.8 | 2,434 | 89.2 | 4,148 | 27,984 | 153,068 |
| Texas | 23.8 | 1,193 | 53.2 | 1,513 | 81.7 | 2,000 |  |  | 25,036 | 97,962 |
| Florida | 23.8 | 1,813 | 44.5 | 903 | 66.4 | 1,074 | 89.2 | 1,848 | 26,098 | 80,625 |
| New York | 22.4 | 11,802 | 43.2 | 4,963 | 72.4 | 1,763 |  |  | 247,566 | 389,010 |
| Pennsylvania | 24.3 | 1,969 | 47.8 | 1,271 | 72.1 | 736 |  |  | 41,307 | 83,431 |
| Illinois | 22.0 | 1,580 | 46.7 | 2,894 | 67.7 | 1,872 |  |  | 33,142 | 133,166 |
| Ohio | 14.6 | 421 | 33.7 | 1,060 | 55.0 | 782 | 73.6 | 681 | 5,836 | 41,149 |
| Georgia | 18.4 | 1,333 | 40.6 | 1,046 | 62.6 | 910 | 82.6 | 907 | 18,611 | 58,441 |
| North Carolina | 19.4 | 432 | 42.1 | 601 | 62.9 | 875 | 81.7 | 1,507 | 6,036 | 47,449 |
| Michigan | 16.9 | 2,367 | 39.5 | 1,098 | 60.1 | 630 | 83.7 | 1,742 | 32,990 | 81,152 |
| New Jersey | 20.0 | 4,375 | 41.0 | 2,675 | 68.7 | 1,003 |  |  | 91,683 | 169,046 |
| Virginia | 25.5 | 514 | 48.7 | 969 | 72.6 | 1,185 |  |  | 10,798 | 55,908 |
| Washington | 23.3 | 583 | 56.8 | 374 | 88.3 | 406 |  |  | 12,234 | 28,499 |
| Arizona | 14.5 | 320 | 37.5 | 370 | 54.3 | 528 | 79.9 | 1,777 | 4,438 | 41,414 |
| Massachusetts | 25.2 | 1,841 | 44.4 | 2,193 | 72.5 | 1,049 |  |  | 38,636 | 106,680 |
| Tennessee | 17.4 | 332 | 46.1 | 552 | 77.0 | 630 |  |  | 6,944 | 31,666 |
| Indiana | 15.2 | 597 | 40.3 | 812 | 66.3 | 548 |  |  | 12,447 | 41,034 |
| Missouri | 12.6 | 317 | 41.2 | 254 | 71.8 | 273 |  |  | 6,563 | 17,627 |
| Maryland | 20.7 | 814 | 43.9 | 1,119 | 66.2 | 1,069 |  |  | 17,056 | 62,946 |
| Wisconsin | 16.3 | 209 | 46.3 | 387 | 71.2 | 521 |  |  | 4,359 | 23,381 |
| Colorado | 16.4 | 451 | 42.3 | 593 | 70.1 | 362 |  |  | 9,409 | 29,496 |
| Minnesota | 19.4 | 109 | 49.7 | 753 | 70.0 | 593 |  |  | 7,916 | 30,507 |
| South Carolina | 20.6 | 243 | 50.2 | 210 | 81.2 | 471 |  |  | 5,093 | 19,222 |
| Alabama | 17.6 | 295 | 45.9 | 308 | 68.1 | 510 |  |  | 6,173 | 23,310 |
| Louisiana | 17.2 | 1,424 | 61.3 | 637 |  |  |  |  | 13,504 | 43,273 |
| Kentucky | 19.6 | 182 | 45.6 | 239 | 76.0 | 260 |  |  | 3,817 | 14,179 |
| Oregon | 11.83 | 98 | 44.1 | 90 | 80.7 | 135 |  |  | 2,028 | 6,670 |
| Oklahoma | 16.6 | 195 | 40.1 | 134 | 59.7 | 143 | 82.3 | 145 | 2,710 | 8,588 |
| Connecticut | 20.1 | 965 | 35.8 | 621 | 59.2 | 532 |  |  | 20,227 | 44,475 |
| Utah | 13.4 | 196 | 37.3 | 234 | 57.1 | 200 |  |  | 2,715 | 16,210 |
| Iowa | 23.3 | 163 | 45.7 | 828 | 68.2 | 493 | 88.8 | 481 | 13,846 | 27,265 |
| Nevada | 16.6 | 249 | 39.4 | 187 | 63.0 | 187 | 86.5 | 309 | 6,071 | 12,894 |
| Arkansas | 21.9 | 151 | 55.5 | 173 | 76.5 | 402 |  |  | 3,170 | 14,869 |
| Mississippi | 12.1 | 200 | 30.7 | 326 | 49.1 | 355 | 69.9 | 488 | 7,310 | 19,104 |
| Kansas | 15.1 | 86 | 42.7 | 350 | 72.2 | 113 |  |  | 8,095 | 11,490 |
| New Mexico | 21.7 | 119 | 46.6 | 201 | 75.9 | 187 |  |  | 2,490 | 10,588 |
| Nebraska | 21.2 | 57 | 45.3 | 487 | 72.2 | 290 |  |  | 11,262 | 17,319 |
| West Virginia | 8.7 | 71 | 30.4 | 27 | 54.8 | 63 |  |  | 941 | 2,244 |
| Idaho | 9.8 | 141 | 42.6 | 45 | 68.9 | 66 |  |  | 1,903 | 3,503 |
| Hawaii | 12.2 | 43 |  |  |  |  |  |  | 0 | 595 |
| New Hampshire | 12.8 | 70 | 40.0 | 110 | 65.6 |  |  |  | 1,436 | 5,405 |
| Maine | 14.8 | 59 | 43.5 | 44 | 63.3 | 69 | 82.7 | 35 | 813 | 2,883 |
| Montana | 2.9 | 34 |  |  |  |  |  |  | 0 | 445 |
| Rhode Island | 16.3 | 351 | 31.6 | 224 | 53.0 | 177 |  |  | 7,321 | 15,776 |
| Delaware | 19.3 | 192 | 37.2 | 267 | 57.6 | 250 |  |  | 2,677 | 9,921 |
| South Dakota | 17.7 | 180 | 43.0 | 151 | 65.4 | 91 |  |  | 2,506 | 5,887 |
| North Dakota | 5.4 | 15 | 30.0 | 57 | 55.8 | 75 |  |  | 1,467 | 3,032 |
| Alaska | 8.29 | 17 | 75.8 | 26 |  |  |  |  | 337 | 863 |
| Vermont | 13.0 | 63 |  |  |  |  |  |  | 0 | 874 |
| Wyoming | 13.7 | 30 | 39.8 | 17 | 61.3 | 17 |  |  | 445 | 957 |
| District of Columbia | 17.7 | 161 | 42.7 | 203 | 66.1 | 101 |  |  | 3,376 | 9,754 |

**TABLE II:** Ranking of the various countries according to the Pandemic Response Index, defined in Eq. (4). *N*_*T*_ is the asymptotic value after all waves have decayed, given in Table I, normalized by the state’s population. *ΔN*_*T*_ is the difference between *N*_*T*_ and the number of cases which were infected by the first major wave *N*_*T*_^*(1)*^ (for the states of Minnesota, Iowa, Mississippi, Kansas, Nebraska, and North Dakota, both the small initial wave and the second wave had been taken into account). The last column contains the PRI_USA_ value (see text for details).
c. *λ*, a parameter introduced to quantify the trend in the number of cases after the first wave, defined as *λ* = 1 + tan *ϕ* (10^3^/*N*_*T*_), where tan *ϕ* is the slope of the linear trend of the 7-day-averaged daily cases in the last 50 days, estimated by ordinary least squares regression (see also [27] for contrasting trends of daily new confirmed infections); the trend can be declining (tan *ϕ* < 0), increasing (tan *ϕ* > 0) or stationary (tan *ϕ* = 0), thus *λ* indicates better performance for those states where the number of cases has been steadily decreasing for this period. The additional factor (10^3^/*N*_*T*_) in the definition of *λ* is required to produce values for this parameter in the range [0.5, 1.5], which can lead to meaningful comparisons.

| USA States | $N_T$ | $N_T$<br>/million. | $\Delta N_T$ | $\Delta N_T$<br>/million | $\lambda$ | $\text{PRI}_{\text{USA}}$ |
| --- | --- | --- | --- | --- | --- | --- |
| California | 153,068 | 3,874 | 125,085 | 3,166 | 1.174 | 4.066 |
| Texas | 97,962 | 3,378 | 72,927 | 2,515 | 1.147 | 4.762 |
| Florida | 80,625 | 3,754 | 54,527 | 2,539 | 1.078 | 5.342 |
| New York | 389,010 | 19,997 | 141,444 | 7,271 | 0.693 | 5.276 |
| Pennsylvania | 83,431 | 6,517 | 42,124 | 3,290 | 0.746 | 6.901 |
| Illinois | 133,166 | 10,509 | 100,025 | 7,893 | 0.848 | 4.587 |
| Ohio | 41,149 | 3,520 | 35,310 | 3,021 | 0.884 | 5.431 |
| Georgia | 58,441 | 5,504 | 39,830 | 3,751 | 0.963 | 5.393 |
| North Carolina | 47,449 | 4,524 | 41,413 | 3,949 | 1.312 | 2.793 |
| Michigan | 81,152 | 8,126 | 48,162 | 4,823 | 0.948 | 5.262 |
| New Jersey | 169,046 | 19,032 | 77,363 | 8,710 | 0.629 | 5.568 |
| Virginia | 55,908 | 6,550 | 45,111 | 5,285 | 1.124 | 3.623 |
| Washington | 28,499 | 3,743 | 16,265 | 2,136 | 1.026 | 6.114 |
| Arizona | 41,414 | 5,690 | 36,976 | 5,080 | 1.314 | 2.267 |
| Massachusetts | 106,680 | 15,351 | 68,044 | 9,791 | 0.654 | 5.403 |
| Tennessee | 31,666 | 4,634 | 24,722 | 3,618 | 1.082 | 4.522 |
| Indiana | 41,034 | 6,095 | 28,587 | 4,246 | 0.878 | 5.603 |
| Missouri | 17,627 | 2,872 | 11,064 | 1,803 | 1.005 | 6.126 |
| Maryland | 62,946 | 10,412 | 45,890 | 7,591 | 0.998 | 3.764 |
| Wisconsin | 23,381 | 4,016 | 19,023 | 3,267 | 1.186 | 3.984 |
| Colorado | 29,496 | 5,122 | 20,087 | 3,488 | 0.813 | 6.189 |
| Minnesota | 30,507 | 5,409 | 12,591 | 2,233 | 1.207 | 5.878 |
| South Carolina | 19,222 | 3,733 | 14,129 | 2,744 | 1.219 | 4.382 |
| Alabama | 23,310 | 4,754 | 17,137 | 3,495 | 1.239 | 3.972 |
| Louisiana | 43,273 | 9,308 | 13,504 | 2,905 | 1.006 | 6.090 |
| Kentucky | 14,179 | 3,174 | 10,363 | 2,320 | 1.022 | 5.453 |
| Oregon | 6,670 | 1,581 | 4,643 | 1,101 | 1.012 | 6.079 |
| Oklahoma | 8,588 | 2,170 | 5,878 | 1,485 | 0.986 | 6.089 |
| Connecticut | 44,475 | 12,474 | 24,248 | 6,801 | 0.654 | 6.178 |
| Utah | 16,210 | 5,056 | 13,495 | 4,209 | 1.199 | 3.496 |
| Iowa | 27,265 | 8,642 | 13,414 | 4,252 | 0.916 | 5.767 |
| Nevada | 12,894 | 4,186 | 6,823 | 2,215 | 1.032 | 6.190 |
| Arkansas | 14,869 | 4,927 | 11,699 | 3,877 | 1.273 | 3.426 |
| Mississippi | 19,104 | 6,419 | 11,794 | 3,963 | 1.084 | 4.913 |
| Kansas | 11,490 | 3,944 | 2,395 | 822 | 0.719 | 8.541 |
| New Mexico | 10,588 | 5,050 | 8,098 | 3,862 | 1.024 | 4.794 |
| Nebraska | 17,319 | 8,953 | 6,057 | 3,131 | 0.933 | 6.282 |
| West Virginia | 2,244 | 1,252 | 1,304 | 728 | 1.037 | 6.661 |
| Idaho | 3,503 | 1,960 | 1,601 | 896 | 1.078 | 7.008 |
| Hawaii | 595 | 480 | 0 | 0 | 0.925 | 9.903 |
| New Hampshire | 5,405 | 3,975 | 3,970 | 2,920 | 0.945 | 5.592 |
| Maine | 2,883 | 2,145 | 2,070 | 1,540 | 1.153 | 5.244 |
| Montana | 445 | 527 | 0 | 0 | 1.151 | 9.880 |
| Rhode Island | 15,776 | 14,892 | 8,456 | 7,982 | 0.615 | 6.062 |
| Delaware | 9,921 | 10,188 | 7,244 | 7,439 | 0.722 | 5.528 |
| South Dakota | 5,887 | 6,655 | 3,381 | 3,822 | 0.915 | 5.848 |
| North Dakota | 3,032 | 3,979 | 1,565 | 2,054 | 0.977 | 6.507 |
| Alaska | 863 | 1,180 | 527 | 720 | 1.290 | 5.680 |
| Vermont | 874 | 1,779 | 0 | 0 | 1.027 | 9.641 |
| Wyoming | 957 | 1,654 | 512 | 885 | 0.927 | 7.137 |
| District of<br>Columbia | 9,754 | 13,821 | 6,379 | 9,039 | 0.816 | 4.511 |

With these three quantities, we then define the “Pandemic Response Index” (PRI) for the USA as:

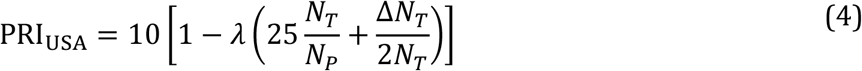

PRI_USA_ values lie in the range from 0 to 10, the higher values corresponding to better performance. This provides a quantitative and objective way of ranking the states according to their performance. The results of this comparison and the relevant numbers that enter in the evaluation of the PRI_USA_ are given in Table II.

## Results

We fit incidence curves corresponding to the infected population data, for each one of the 50 USA states and the District of Columbia. We fit seven-day moving averages of the reported daily data, with data up to June 11, 2020. For each US state we estimate the number of waves in which the infected population curve can be analyzed, the values of the model parameters of each wave, and the expected number of cases for the first major wave, *N*_*T*_^*(1)*^, and for all waves, *N*_*T*_. Table I presents the model parameters for each state.

The response of each state can be classified in three distinct classes. The first class comprises those that exhibit a large initial wave followed by several smaller ones. This class can be divided in two subclasses: the first comprises states of smaller populations and population densities, and correspondingly small number of cases, containing the epidemic in a *single* wave (Hawaii, Vermont, Montana). Initially, the State of Alaska was also included in this subclass, as a single wave state. However, as will be shown in Fig. 6, after the decay of the first wave a second wave, non-overlapping with the first, of higher intensity, was generated. However the states of Hawaii, Vermont, and Montana have recently exhibited additional cases, which may become a second wave, as in the case of Alaska; as of June 11, 2020, only the State of Hawaii can be qualified in this subclass as the increasing number of cases of the State of Vermont and of the State of Montana classify them to the second class and the third class, respectively. The second subclass comprises states with high numbers of cases, large population sizes and urban centers, mostly implementing rather strict measures, exhibiting multiple waves of the epidemic. States in this subclass include New York, Pennsylvania, Illinois, Ohio, Massachusetts, Michigan, Connecticut, New Jersey, Rhode Island, Colorado, Indiana, Kansas, Nebraska, New Hampshire, Delaware, Iowa, Wyoming, South Dakota, and the District of Columbia. In these states, an initial large wave is followed by a series of waves of declining strengths, trending toward the containment of the Covid-19 disease (in some states there is an initial smaller wave followed by the larger one). This behavior signifies the imposition of social distancing measures obeyed by the citizens, an effect which is corroborated by recent Bayesian inference studies [14] and agent-based simulations [15].

The second class comprises states in which an initial large wave is followed by waves that do not systematically decline or increase, leading to an averaged plateau of reported cases. New sub-epidemics seem to appear and the reported cases in these states are not exhibiting a declining trend but they are not exhibiting an increasing trend either. States in this “stationary” (endemic-type wave) class include Georgia, Washington, Maryland, Missouri, Louisiana, Oklahoma, New Mexico, West Virginia, North Dakota, Kentucky, Oregon, Nevada, and Vermont.

The third class comprises states exhibiting an *increasing* trend of reported cases and includes the states of California, Texas, Florida, North Carolina, Virginia, Arizona, Tennessee, Wisconsin, Minnesota, South Carolina, Alabama, Utah, Arkansas, Mississippi, Maine, Montana, Idaho, and Alaska.

Of the 50 states and the District of Columbia, 20 belong to the first class, exhibiting declining trends, 13 belong to the second class, exhibiting stationary behavior, and 18 belong to the third class, exhibiting increasing trends. The states are classified to each class according to their λ values (detailed ranking is depicted in Fig. 8). Overall, as of June 11, 2020, only 40% of the states show pandemic response leading to clearly declining trends in the number of cases. A color map of the USA depicting the states belonging to each one of these classes is presented below, with the 3 classes color-coded.

In the following Figures (Fig.s 2-6), we present representative results obtained by the multiple-wave FSIR model for selected states that can be accurately fitted by 4 waves (California, Florida, Georgia, Arizona), 3 waves (Illinois, Texas, New York, Pennsylvania), and two waves (Alaska, Idaho). These states also represent the three distinct classes of epidemic response.

**Figure 1:**
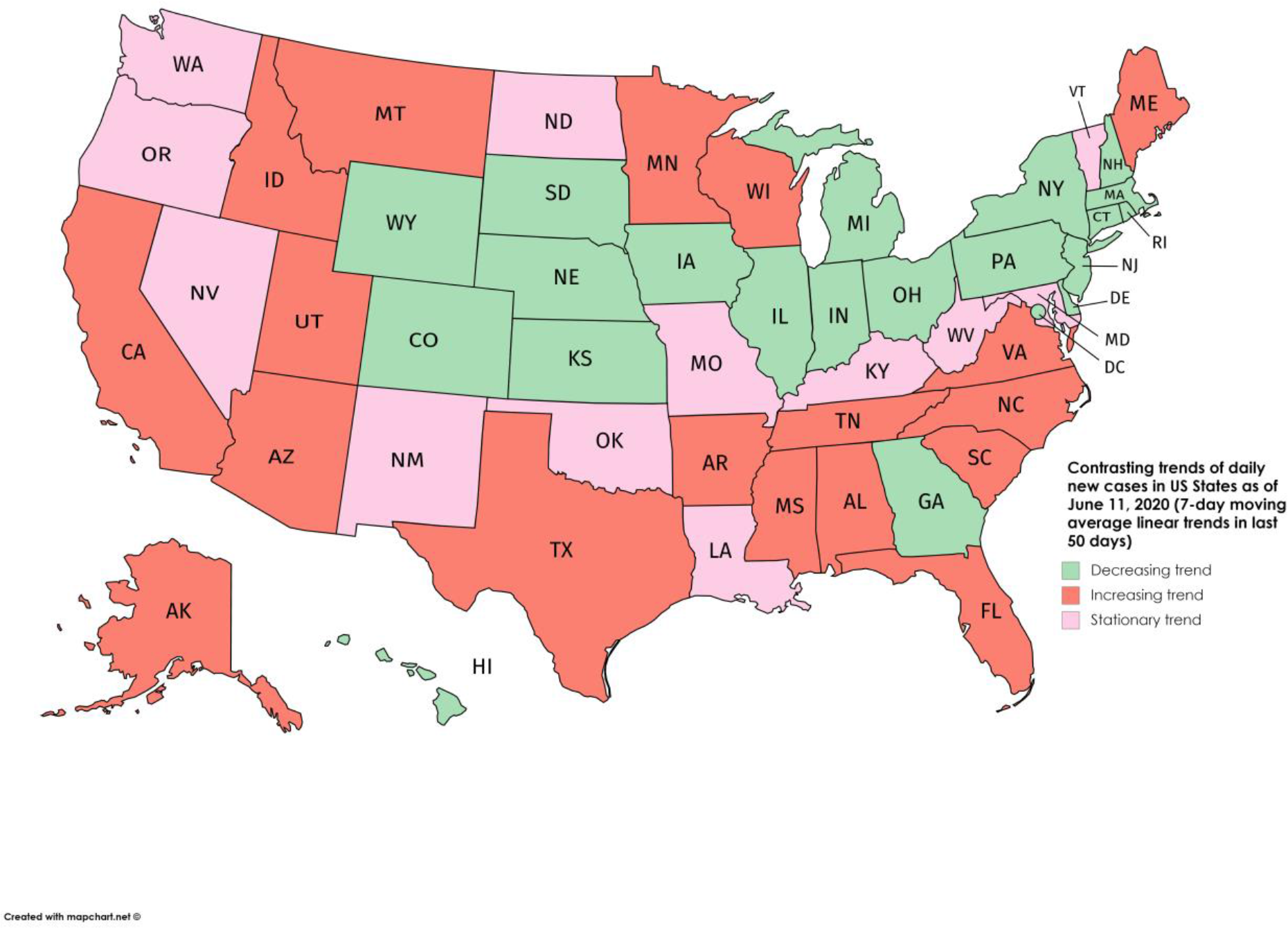
A color-coded map classifying the epidemic trends in the 50 USA states and the District of Columbia, according to their *λ* values (see Fig. 8 and text for details).

**Figure 2:**
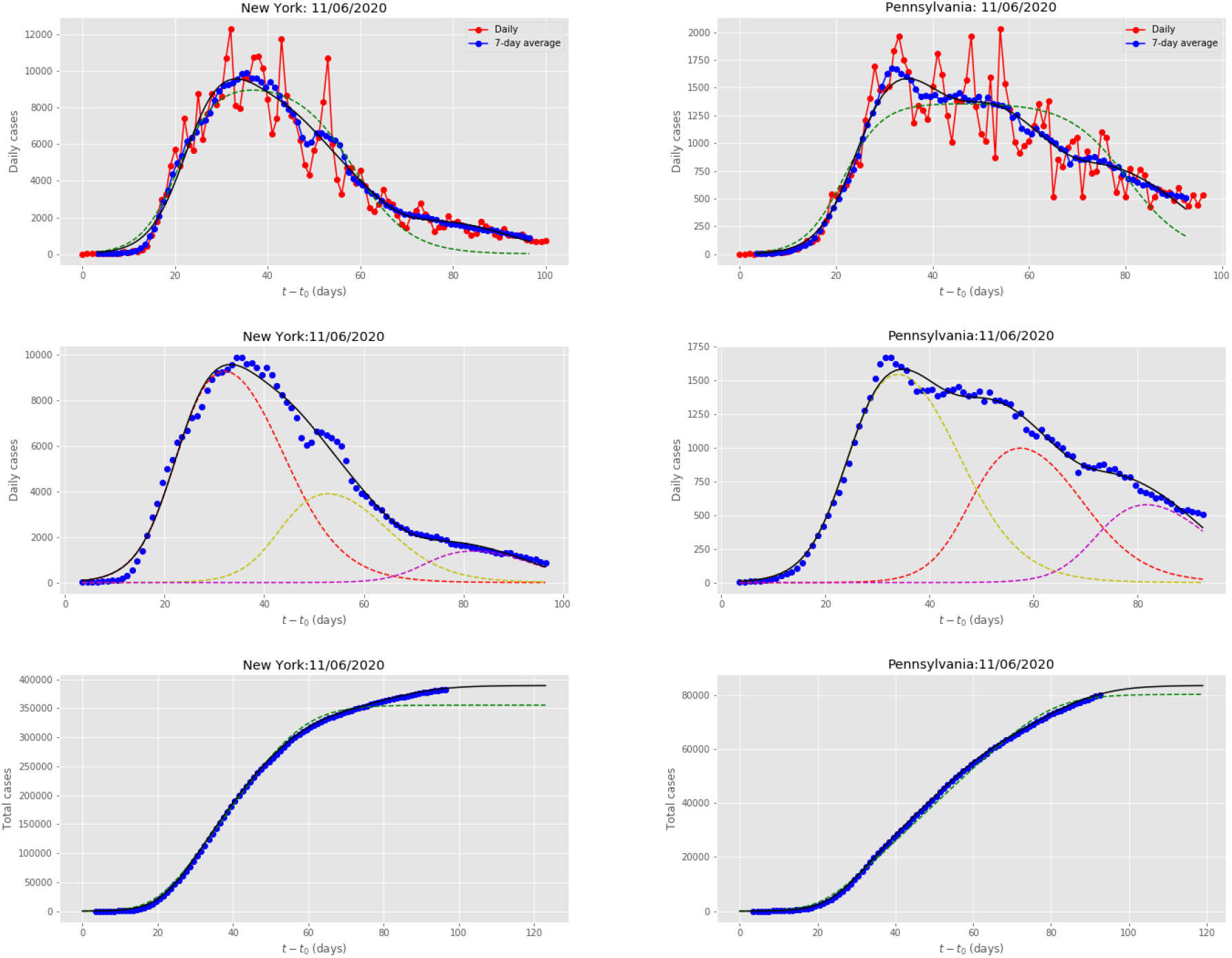
Results for New York and Pennsylvania, obtained by fitting the multiple-wave FSIR model with data up to June 11, 2020. **Top row**: Red dots are the daily data reported in Ref. [23]. The blue dots are seven-day running averages of the daily data. The green-dashed line is the fit by the single-wave FSIR model. The black solid line is the 3-wave fit by the multiple-wave FSIR model. **Middle row**: Decomposition of the seven-day moving average data (blue dots) in 3 waves, for each state. The black line represents the superposition of the multiple waves. The fit is in excellent agreement with the actual data. **Bottom row**: Blue dots are cumulative daily data (moving averages). The black line is the fit by the multiple-wave FSIR model, and it is essentially indistinguishable from the actual data. The green-dashed line is the fit of the single-wave FSIR model, which clearly underfits the actual data.

Fig. 2 presents the multiple-wave fit for New York, which has been the most heavily impacted state by the Covid-19 pandemic, in absolute numbers. An initial wave was followed by two declining waves, manifesting New York’s imposition of strict intervention measures. There is an excellent agreement between the 3-wave fit and the actual data, in both daily (averaged over 7-day periods) and cumulative data. As can be seen, the single-wave fit of the data, depicted by the green-dashed lines, significantly underfits the data. Fig. 2 also presents the multiple-wave fit for Pennsylvania, fitted by 3 waves. Here again, an initial large wave was followed by two declining sub-epidemics, signifying the imposition of distancing measures. The agreement between the 3-wave fit and the actual data, in both daily and cumulative data, is equally good, and the single wave fit of the data significantly underfits the actual data. New York and Pennsylvania are representative examples of states where the initial major wave is followed by several waves of *declining* strength, suggesting that, despite the initial large impact, the states are well on the way to eventually contain the epidemic.

Fig. 3 presents the multiple wave fits for Texas and Illinois. The incidence curves of both states were fitted by 3 waves. Texas, the 2^nd^ most populous state, had a low total number of cases, the lowest among the 10 most populous states in USA; Illinois is the 6^th^ most populous state. Both states have exhibited two waves of *increasing* intensity. However, Illinois exhibited a subsequent wave of declining strength whereas Texas exhibited one with increasing strength. It remains to be determined if this increasing trend will continue. Since there is no clear trend of the infected population in Texas to getting over its intensity peak, the single-wave fit (depicted by the dashed green line) predicts a linear increase of the total number of expected cases. For the State of Illinois, the 3-wave fit, for the cumulative cases, estimates a final plateau signifying the containment of the epidemic after the third wave, assuming of course that no more waves of increased strength materialize.

**Figure 3:**
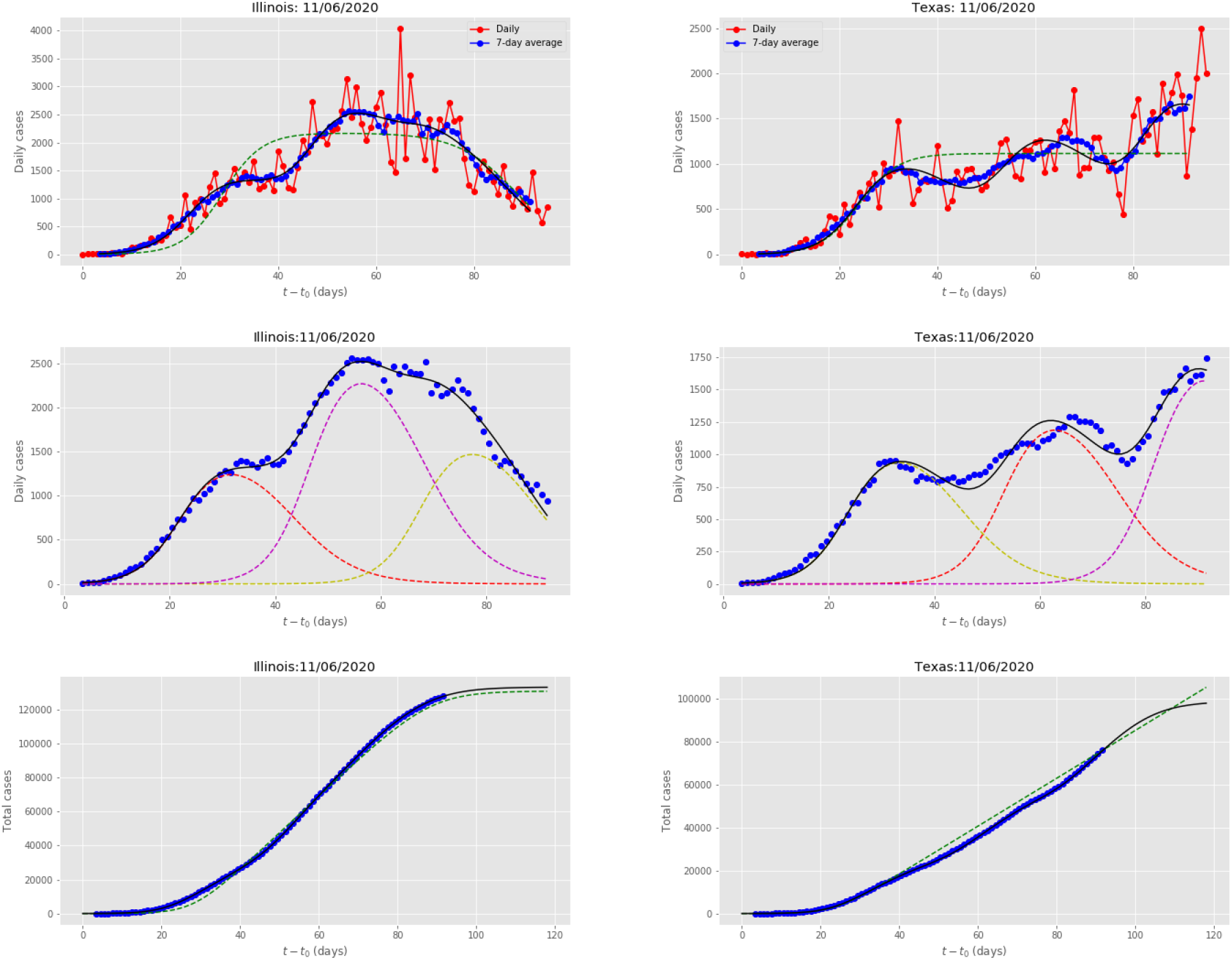
Results for Illinois and Texas, obtained by fitting the multiple-wave FSIR model with data up to June 11, 2020. The meaning of symbols is the same as in Fig. 2.

Fig. 4 presents the multiple wave fits for Florida and Georgia. These states implemented measures to stop the disease’s transmission and impact, which were deemed not aggressive and fast enough; furthermore, they were among the states that “re-opened” rather fast. The incidence curves were fitted by 4 waves. Florida exhibited an initial large wave followed by two sub-epidemics of declining strength. However, the declining trend did not continue but was intercepted by a sub-epidemic of increased strength. Georgia has exhibited a series of sub-epidemics generating a plateau of reported cases, which can be classified as a stationary (endemic) wave according to the classification scheme of [13]; it remains to be determined if this endemic-type wave will be stationary or temporary.

**Figure 4:**
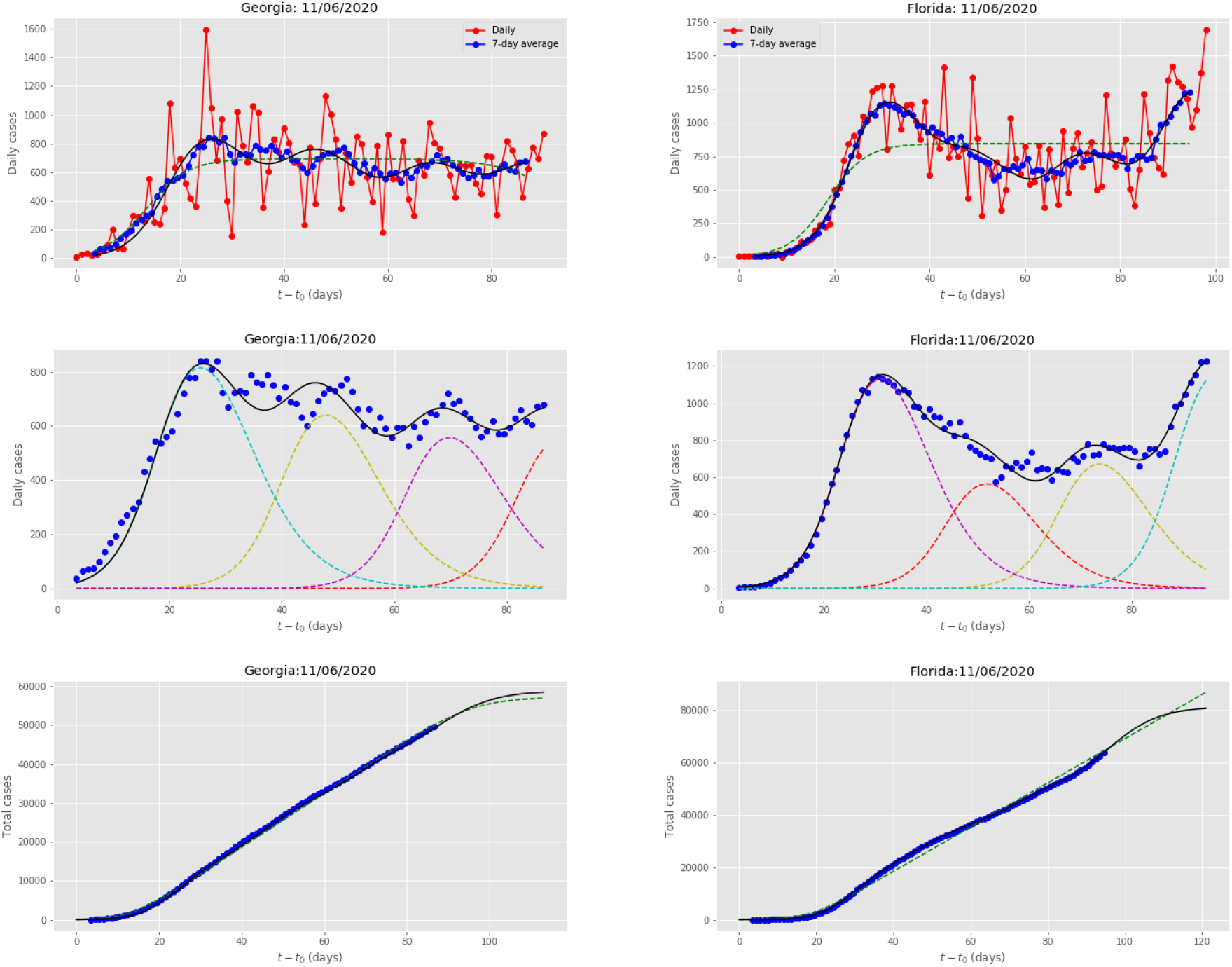
Results for Georgia and Florida, obtained by fitting the multiple-wave FSIR model with data up to June 11, 2020. The meaning of symbols is the same as in Fig. 2.

Fig. 5 presents the multiple-wave fits for California and Arizona. The states’ incidence curves were fitted by 4 waves. Both states exhibited sub-epidemics with *increasing* strengths. The states had taken intervention measures rather fast. However, the disease was spreading in increasing intensity waves in both states. Recent results on the introduction and spread of the virus in Arizona [19], based on sequencing viral genomes from clinical samples, revealed a minimum of 9 distinct introductions throughout February and March 2020. Since there is no clear trend as of June 11, 2020, of each state getting over its intensity peak, the single-wave fit predicts a linear increase of the total number of expected cases. The 4-wave fit estimates a plateau of the total number of cases after the fourth wave, assuming that additional waves do not materialize.

**Figure 5:**
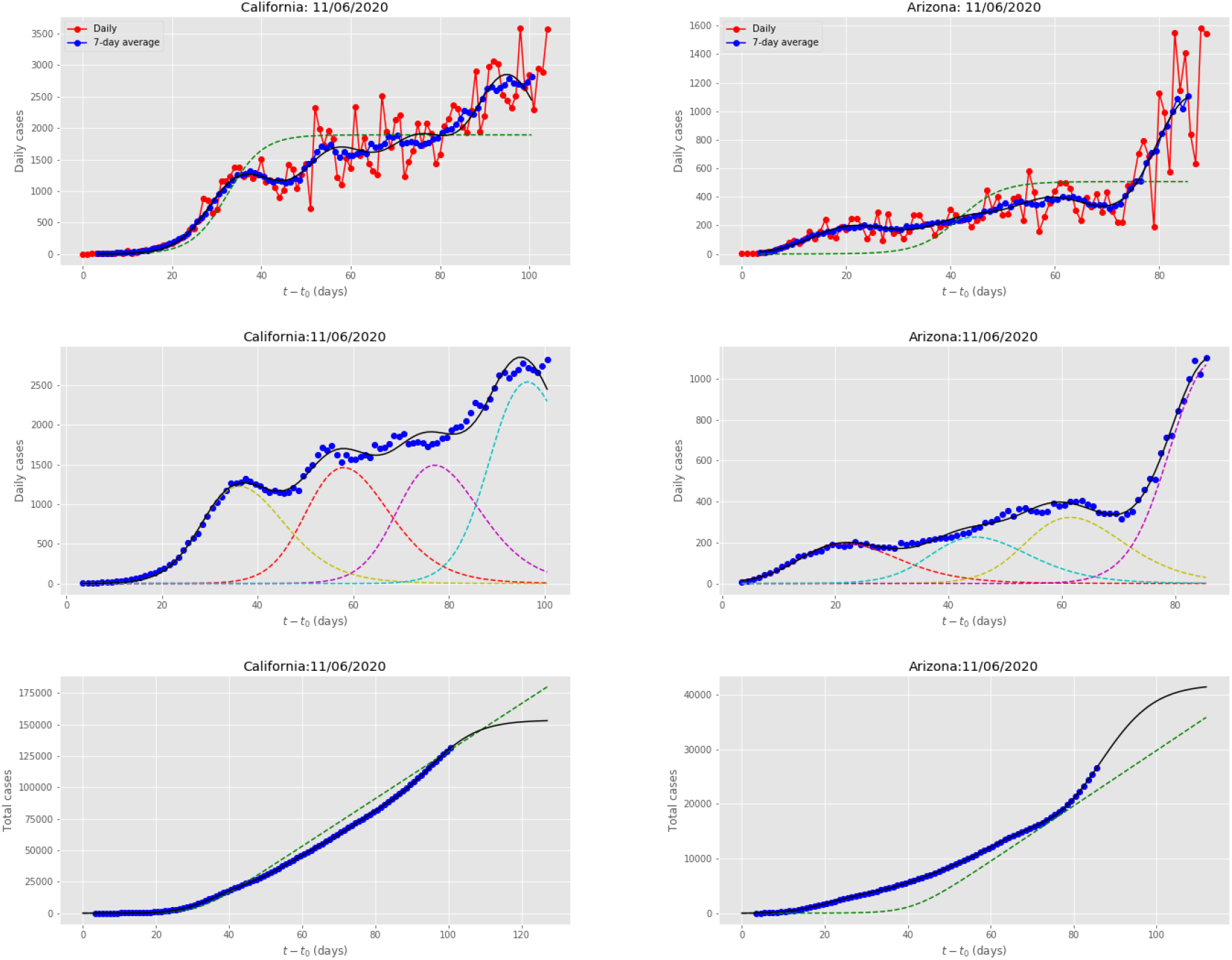
Results for California and Arizona, obtained by fitting the multiple-wave FSIR model with data up to June 11, 2020. The meaning of symbols is the same as in Fig. 2.

California, Texas, and Arizona are representative examples of states where the initial major wave is followed by several waves of *increasing* strength, suggesting that, despite the initial impact, the states were not able to contain the spread of the disease. On the other hand, Georgia is representative example of states where the initial wave is followed by sub-epidemics of almost stationary intensities, producing essentially an endemic-type wave.

Finally, Fig. 6 presents the multiple-wave fits for Idaho and Alaska. These responses can be considered as “outlier” cases. After an initial large wave, Idaho’s infected population exhibited a stationary (endemic) wave but with an increasing trend. The incidence curve of Idaho was fitted by 3 waves in total (the endemic component of Idaho’s infected curve was modeled by fitting two waves). Fig 6 also presents a 2-wave fit for Alaska, which shows a first, single, wave, followed at a later time, by a second wave of a higher peak, non-overlapping with the former wave, a feature that can categorize the latter wave as the “second Covid-19 wave”, in the meaning usually assigned to this term by the media [28, 29].

**Figure 6:**
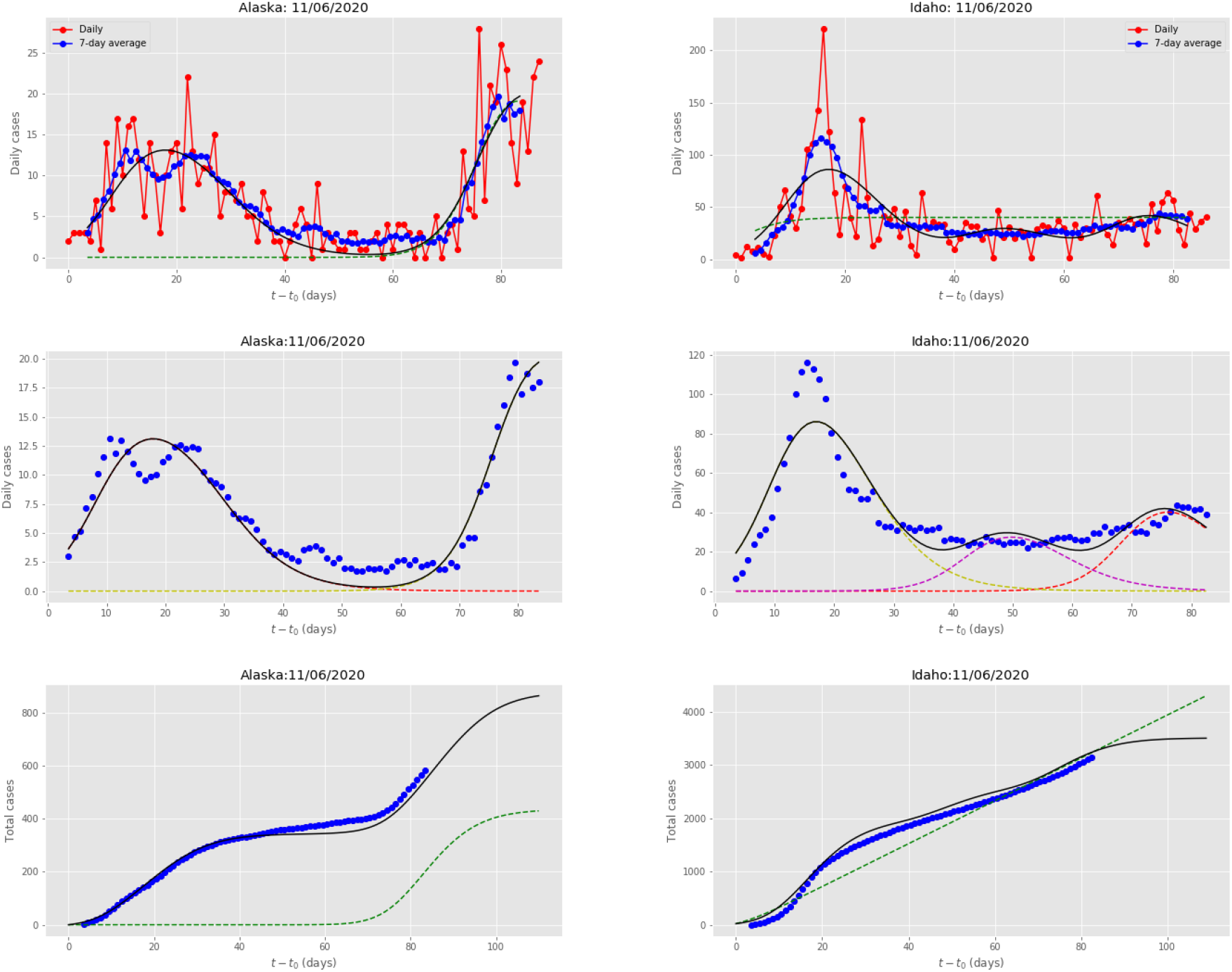
Results for Alaska and Idaho, obtained by fitting the multiple-wave FSIR model with data up to June 11, 2020. The meaning of symbols is the same as in Fig. 2.

We can quantify each state’s response with the help of the Pandemic Response Index, Eq. (4), and make comparisons, which may be useful for contributing to the evaluation of different underlying factors and different policies followed. Table II presents (a) the total number of infections as given by the quantity *N*_*T*_, the number of infections over the state’s population, *N*_*T*_*/N*_*P*_, as well as *N*_*T*_ per million population; (b) the number of infections that correspond to all the waves except for the first major one, Δ*N*_*T*_ = *N*_*T*_ *– N*_*T*_^*(1)*^; (c) the PRI_USA_ value, based on the values of the parameters *N*_*T*_, *N*_*T*_*/N*_*P*_, and Δ*N*_*T*_ / 2*N*_*T*_.

Hawaii, Montana, and Vermont show the highest PRI_USA_ values (9.90, 9.88, and 9.64, respectively) because of their small number of cases and the essentially single wave epidemic curve, for which Δ*N*_*T*_ =0. For 16 US States, the PRI value is less than 5. The PRI_USA_ average of all US States and the District of Columbia is 5.602, whereas the population-weighted PRI_USA_ average is 5.048. We note that this average value should not be taken as the country’s Covid-19 RPI value, when compared to other countries, because a different scale applies to different sets depending on the range of *N*_*T*_ values of the members of the comparison set.

In Fig. 7, we present rankings of USA sates according to *N*_*T*_ (left panel) and to *ΔN*_*T*_ (right panel) values. Both rankings contain useful information but do not capture the more complete picture needed to evaluate each state’s response. In Fig. 8, we present the ranking of USA states according to the values of *λ* (left panel), which provides a measure of whether the number of cases has been rising (*λ* > 1), stationary (*λ* ≈ 1), or falling (*λ* < 1). We also present the ranking according to values of the *PRI*_*USA*_ index, defined in Eq. 4, which give a more complete picture of the overall performance, including the values of *N*_*T*_, *ΔN*_*T*_, and *λ*.

**Figure 7:**
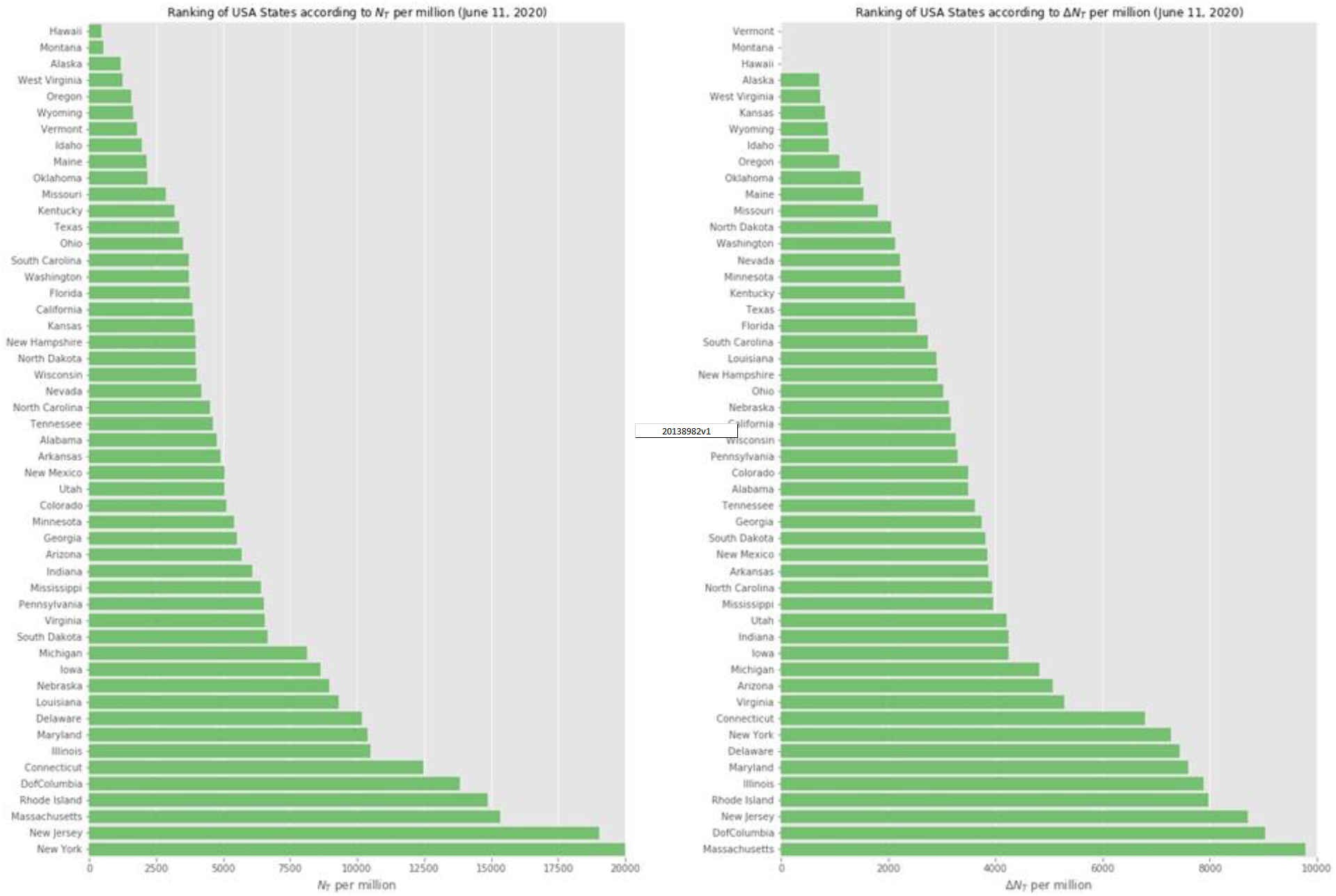
Ranking of USA States as of June 11, 2020. **Left panel**: Ranking according to the values of *N*_*T*_ per million, the expected total number of cases per million population. **Right panel**: Ranking according to the *ΔN*_*T*_ values (see text for details).

**Figure 8:**
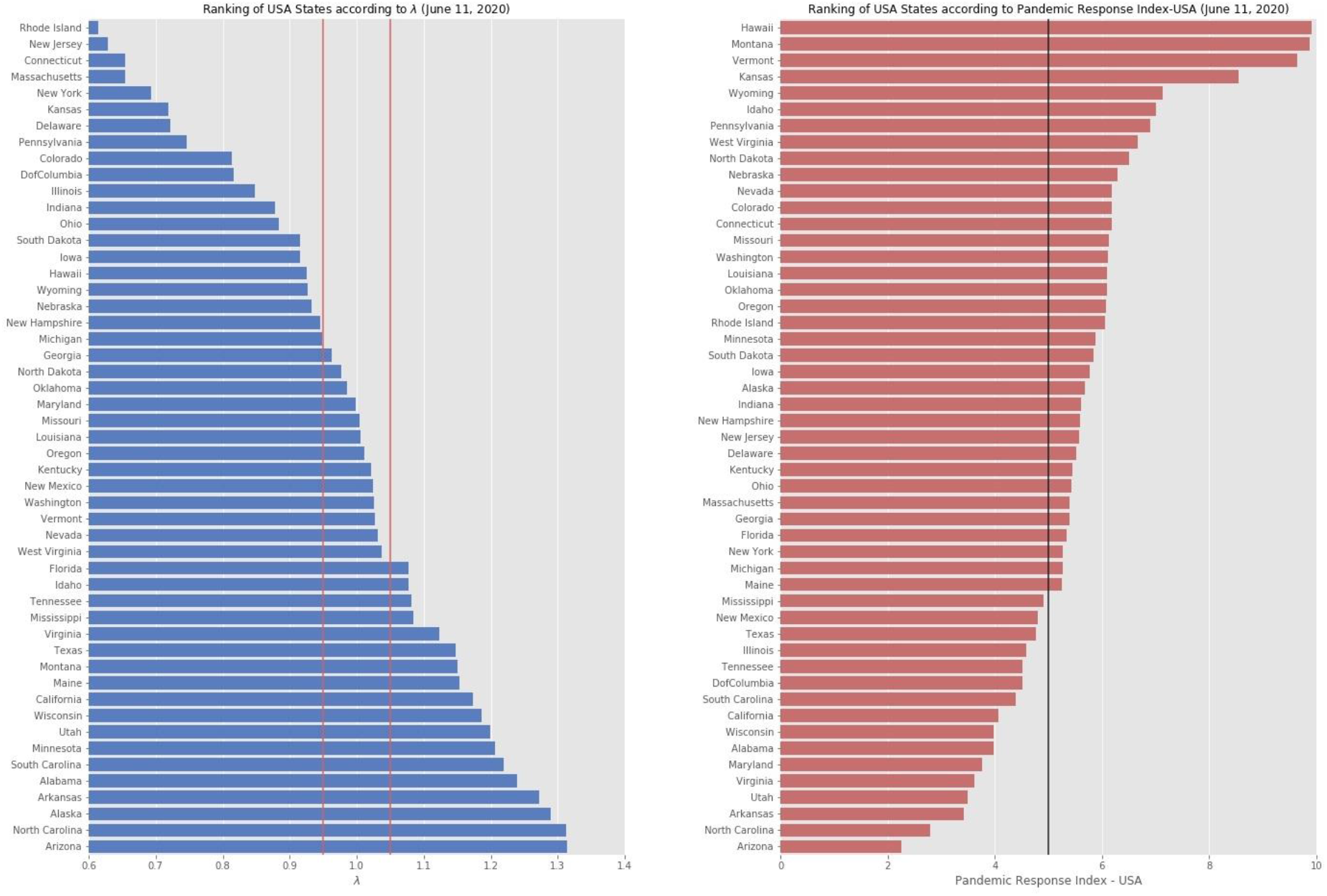
Ranking of USA States as of June 11, 2020. **Left panel**: Ranking according to the values of *λ*, which provides a measure of whether the number of cases has been rising (*λ* > 1), are stationary (*λ* ≈ 1), or falling (*λ* < 1); see text for details. The two vertical red lines delineate the *λ* ≈ 1 range (0.95 < λ < 1.05). States with clearly declining trends have λ < 0.95, whereas states with clearly increasing trends have λ > 1.05. **Right panel**: Ranking according to *PRI*_*USA*_ index. The greater the value of PRI, the better the pandemic response of the state is.

## Discussion

### Principal findings

Reported cases of Covid-19 infections in the USA states show features that are both common and regular, which we interpret as successive waves of transmission (sub-epidemics, outbreaks). We present evidence for this interpretation by analyzing the reported data from all 50 states and the District of Columbia. Based on this analysis, it is possible to infer to what extent the imposition of social-distancing measures had slowed the spread of the disease. This analysis provides an estimate of how much lower the number of infections could have been, if early and strict intervention measures had been taken to stop the spread at the first wave, and assigns a Pandemic Response Index value to each state’s overall response.

### Comparison with prior work

Agent-based simulations encompassing strong social-distancing measures have shown the emergence of epidemic trajectories with multiple wave structures. Recent research works have focused on interpreting the wave features appearing on the reported incidence curves by “change points” [14] resulting from change in the epidemiological parameters after the imposition of interventions or by decomposing the epidemic trajectory in multiple waves.

A recently proposed model, the multiple-wave FSIR model can identify multiple waves, specifying *each one* by only three parameters, *t*_1_, Δ*t* and *N*^’^, all of which are obtained by directly fitting the reported data of daily populations of infected individuals. Each of these parameters can be assigned a physical meaning, which help quantify certain generally held views [22]. Moreover, the quantitative picture that emerges from the values of these parameters produces a rather accurate picture of the severity of the epidemic in the various countries (or states, or counties, or cities), and helps to determine the effect of the intervention measures if and when any were taken.

The multiple-wave model addresses a limitation of models that assume a single wave epidemic. These single-wave models, such as the models employing logistic functions, provide the extrapolation to future cases of infection as only a *lower limit*; this point has been discussed in an elegant mathematical analysis of the data by Fokas *et al*. [30], highlighting the need for the inclusion of nonlinear terms in the underlying differential equations to capture the slow rate of the infected population decay. This is evident in the countries that have long passed the peak of the reported cases: the tail does not asymptote to a constant value, as the sigmoid (logistic) model predicts, but the number actually keeps growing at a slow rate. The multiple-wave FSIR mitigates this limitation of the original FSIR model: by modeling more accurately the wavy behavior of the infected population curve it can provide a better fit to the daily data and to the cumulative actual data, and a better estimate to the cumulative number of cases (*N*_*T*_), as can be seen in all the cases we examined, see Fig.s 2 -6.

### Limitations

The multiple-wave FSIR model we have used in the present study may suffer from a limitation relating to the fact that in many cases, when Δ*t* is estimated as an adjustable parameter, it tends to provide an aggregate fit, that is, an initial large sub-epidemic tends to be followed by a longer in time and smaller in peak intensity averaged wave, which is the sum of smaller sub-epidemics. This wave can be characterized as a temporary endemic wave according to the taxonomy of [13]. In order to improve the resolution of the model and enable it to specify the underlying smaller sub-epidemics, an epidemiologically reasonable value for Δ*t* is necessary (see Section II for details in choosing the Covid-19 epidemiologically consistent value of Δ*t* used in our computations). Furthermore, caution must be exercised in interpreting the sub-epidemics, because they may constitute a superposition of even smaller ones, as in the case of states with significant urban centers in their jurisdiction comprising counties and cities with varied response to the epidemic; a study of multiple-wave decomposition of reported US cases per county and city is currently under way by the authors. In addition, extrapolated values such as the total number of cases, *N*_*T*_, depend on the number of waves, which can produce temporary or stationary “endemic” waves, leading to inaccurate extrapolated values of certain parameters.

Considerable caution should be taken when comparing PRI values outside the set of states (or countries, or cities, or counties) for which the PRI index was calibrated. As mentioned above, RPI is not a “universal” index and it has to be appropriate calibrated for a specific set in order to make meaningful comparisons among the members of the set.

## Conclusions

Multiple waves of transmission during infectious disease epidemics represent a major public health challenge. The analysis of reported data from all 50 states and the District of Columbia in USA support the hypothesis that the Covid-19 pandemic can be successfully modeled as a series of epidemic waves. Based on the model’s results, as of June 11, 2020, only 19 states and the District of Columbia show epidemic waves with declining strengths, leading to the disease’s containment. This is just 40% of the total number of the States. On the other hand, 18 states exhibit waves of increasing strengths, and 13 states exhibit stationary behavior, which can be interpreted as “endemic” wave. Based on the model’s results, we have computed Pandemic Response Index values for each state, thus providing a finer tool for evaluating each state’s Covid-19 response performance.

## Data Availability

All multiple-wave plots of US states will be available as part of the electronic supplementary material.

## Acknowledgements

We are grateful to Eleni Angelaki for her contribution in coding certain parts of the software used.

